# Variation in prescription duration for long term conditions: a cohort study in English NHS primary care using OpenPrescribing

**DOI:** 10.1101/2024.05.28.24308058

**Authors:** Brian MacKenna, Andrew D Brown, Rich Croker, Alex J Walker, Apostolos Tsiachristas, Dave Evans, Peter Inglesby, Ben Goldacre, Seb Bacon, Helen J Curtis

## Abstract

**Background:** Many patients receive routine medications for long-term conditions (LTCs). Doctors typically issue repeat prescriptions in one to three month durations, but England currently has no national guidance on the optimal duration.

**Methods:** We calculated the duration of prescriptions for common LTCs in England over a 12-month period (December 2018-November 2019). We assessed the level of variation between regional clinical commissioning groups (CCGs) and determine practice factors associated with different durations.

**Results:** Of the common medications included, 28-day (one-monthly) prescriptions accounted for 48.5% (2.5 billion) tablets/capsules issued. There was very wide regional variation in the proportion of 28-day prescriptions (7.2% to 95.0%). Practice dispensing status was the most likely predictor of prescription duration. The proportion of patients with LTCs and the electronic health record software used by a practice were also associated with prescription duration.

**Conclusions:** One month prescription durations are common for patients taking medicines routinely for long term conditions, particularly in dispensing practices. Electronic health record configurations offer an opportunity to implement and evaluate new policies on repeat prescription duration in England.

## Background

General practices in England issue 1.1 billion prescriptions every year,[1] two thirds of which are estimated to be repeat prescriptions, commonly for long-term conditions (LTCs).[2] However, NHS England does not issue national guidance on the duration of prescriptions, and doctors are recommended to select a “clinically appropriate” duration.[3] There was a common understanding that 28-day (one month) durations were preferred, based largely upon the principle of minimising wastage from unused medication.[4] However, Scotland and Northern Ireland generally issue a greater proportion of longer prescriptions than England [4], and since 2022, the NHS in Wales is recommending two-month rather than one-month prescriptions where appropriate.[5] Based upon a systematic review and economic modelling, it has been suggested that NHS England should recommend three-monthly prescriptions for LTCs.[6] This recommendation was based on the expectation for increased adherence with medication, reduced inconvenience for patients, and net saved costs in terms of staff time and pharmacy dispensing fees that are likely to outweigh any potential costs of unused medications.[7–10]

Our OpenPrescribing.net service is a publicly funded and openly accessible explorer for all prescriptions dispensed in primary care in England. It was launched in 2015, and has 20,000 unique users every month, including doctors, pharmacists and patients. It displays numerous predefined standard measures for safety, cost, and effectiveness for every practice in England. This includes a measure which displays the proportion prescribed as seven-day durations for a subset of products, but does not yet describe longer durations.[11]

We therefore set out to: describe current prescription durations for common LTCs in England, explore and visualise geographical variation, and identify practice factors associated with shorter prescribing duration to inform policy making.

## Methods

### Study design

Retrospective observational cohort study using prescribing data from all English NHS general practices over one year, December 2018 - November 2019.

### Data sources

We extracted prescribing data from the OpenPrescribing.net database. This imports publicly accessible files published by the NHS Business Services Authority, containing data on prescriptions dispensed for each month, for every prescribing organisation in NHS primary care in England since mid-2010.[12] These monthly datasets contain one row for each strength and formulation of a medication (e.g. atorvastatin 20mg tablets), per organisation. Columns describe ‘items’ (the number of times each medication was issued on prescriptions), ‘total quantity’ (total unit doses issued across all items, e.g. number of tablets), and cost. Crucially for the present analysis, these data are split according to the ‘quantity per item’, the amount of medication dispensed per prescription (e.g. 28 tablets, 56 tablets, etc). These data are sourced from community pharmacy claims and therefore contain all items that were dispensed.

Data on which EHRs are deployed in each general practice were extracted from a monthly file, circulated by NHS Digital to interested parties and available on request [13] (now published annually.[14]) Data on GP practice characteristics were obtained from Public Health England.[15]

### Basket of medicines

We extracted all available prescribing data from mid-2010 to November 2019, the latest data available at the time of analysis prior to the COVID pandemic. We identified a basket of medicines commonly prescribed for LTCs suitable for analysis, using a prior method used for seven-day prescribing measures on OpenPrescribing.[11] Firstly, we identified the top 50 most commonly prescribed tablets/capsules by chemical substance. Since the data does not include dosage instructions e.g. “take two tablets twice a day”, prescribing duration cannot be accurately calculated for medications with mixed dosing regimens. So, we limited the basket of medicines to those nearly exclusively prescribed “once daily” in tablet or capsule form, based upon BNF dosing and clinical experience of two senior pharmacists, resulting in a basket of five medicines (Table 1).

**Table 1.**
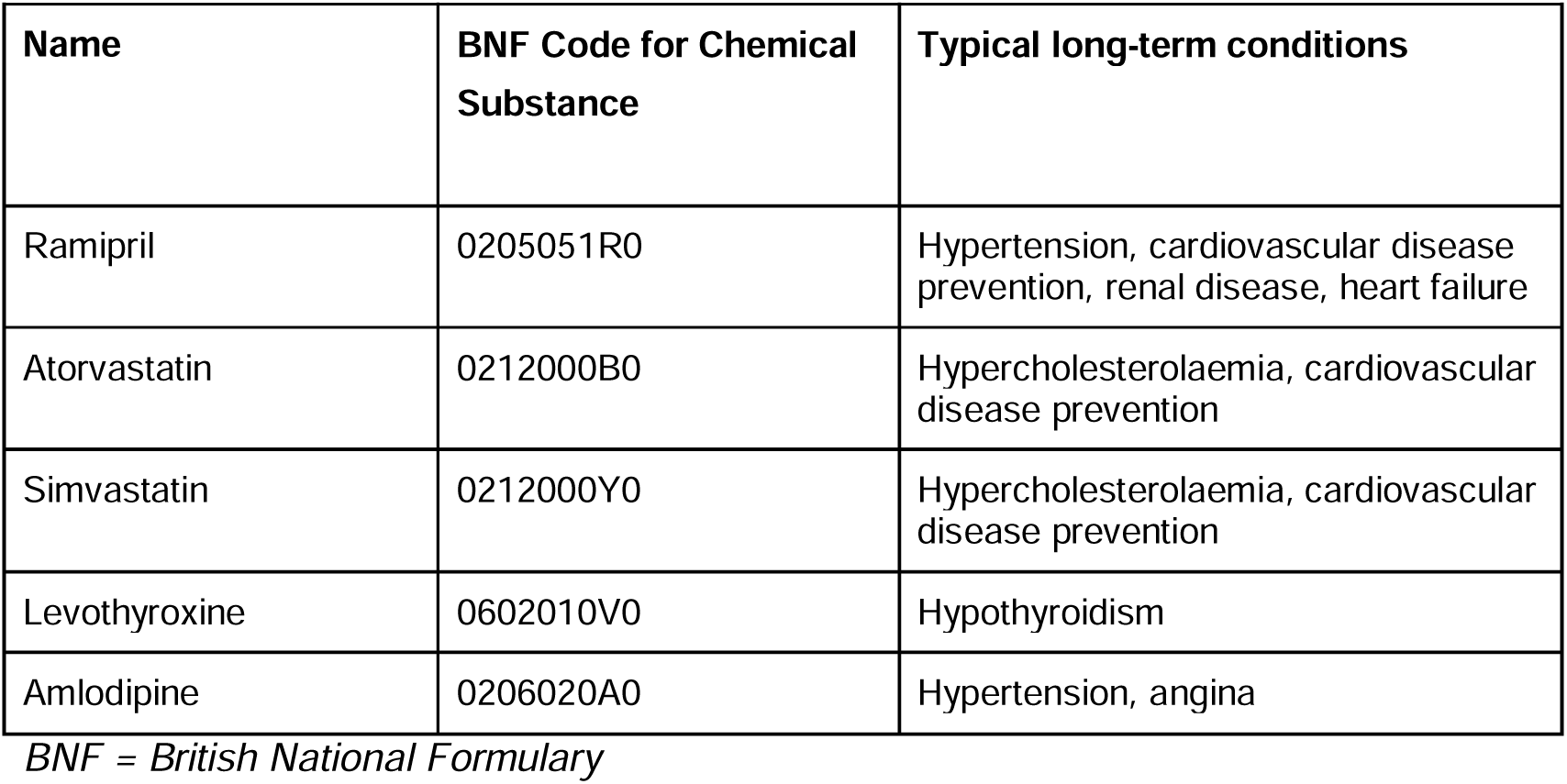
Medications included in analysis. Only tablet/capsule forms are included.

| Name | BNF Code for Chemical Substance | Typical long-term conditions |
| --- | --- | --- |
| Ramipril | 0205051R0 | Hypertension, cardiovascular disease prevention, renal disease, heart failure |
| Atorvastatin | 0212000B0 | Hypercholesterolaemia, cardiovascular disease prevention |
| Simvastatin | 0212000Y0 | Hypercholesterolaemia, cardiovascular disease prevention |
| Levothyroxine | 0602010V0 | Hypothyroidism |
| Amlodipine | 0206020A0 | Hypertension, angina |

BNF = British National Formulary

### Data processing

We extracted all available data covering December 2018 to November 2019 (latest available 12-month period at the time of analysis) for the basket of medicines, limited to those in tablet, capsule, caplet or equivalent forms by those containing the string “Tab” or “Cap” in the medication name. We included only standard general practices, excluding other organisations such as walk-in centres, prisons and hospitals according to the NHS Digital dataset of practice characteristics. We excluded practices that had not prescribed any of the five selected medications. We grouped practices into their parent Clinical Commissioning Groups (CCGs) for regional analyses.

### Prescribing durations

We summed the items issued per duration, then plotted the totals on a histogram. For further analyses we filtered the data to the most common quantity per item values, which were multiples of 28 days: 28 (approximately one month), 56 (approximately two months), or 84 (approximately three months) tablets/capsules. We excluded seven-day prescribing as this is more likely to be done with clinical justification or provision of “dosette box” [11], and therefore not easily amenable to switching to longer durations. We calculated the proportion of medicines issued for each duration, using total *quantity* (number of tablets/capsules), as the number of *items* cannot be directly compared (i.e. 28-day prescriptions require ∼12 prescriptions [items] per year while 84-day prescriptions require only ∼4).

### Geographical variation at CCG level across England

We calculated the proportion of prescriptions issued for each duration (28, 56 and 84 days) out of the total issued for 28, 56 or 84 days, across CCGs in England. We display this geographical variation on choropleth maps.

### Factors associated with prescribing durations

To examine practice factors associated with 28-day prescribing, we used a mixed-effects Poisson regression model. For dependent variable, we defined the “28-day prescribing proportion” as the proportion of prescriptions which were issued for 28 days out of the total issued for 28, 56 or 84 days. Furthermore, we used publicly available data specify the following variables: proportion of GP registered population aged over 65, proportion of patients with a LTC, being a ‘dispensing practice’ with an in-house pharmacy service (yes or no), and the electronic health record (EHR) software system (EMIS, TPP SystmOne, Vision, or MicroTest). These variables were previously shown to be associated with variation in prescribing, and therefore they were included in the regression analysis as independent variables with fixed effect coefficients. Random intercepts were specified in the model to accommodate variation between CCGs. Continuous variables were grouped a priori into quintiles for the analysis. Practices with missing data for a particular variable were not included in models containing that variable. From the resulting model, incidence rate ratios were calculated, with corresponding 95% CIs. The level of missing data was determined and reported for each variable.

For factors with a significant association, we plotted a histogram of practice counts versus 28-day prescribing proportion (unadjusted figures).

### EHR System User-Interface Evaluation

One senior pharmacist tested the interface of the two most widely used EHRs in the study period (EMIS and SystmOne) by issuing prescriptions to a test-patient and observing the prompts. We also contacted the vendors of all four EHRs to enquire about their default options for prescribing duration.

### Software and Reproducibility

Data management was performed using Python 3 and Google BigQuery, with analysis carried out using Stata 13.2 / Python 3. Data and all code for data management and analysis are archived online.[16]

### Patient and Public Involvement

Our website OpenPrescribing.net, is an openly accessible data explorer for all NHS England primary care prescribing data, which receives a large volume of user feedback from professionals, patients and the public. This feedback is used to refine and prioritise our informatics tools and research activities. Patients were not formally involved in developing this specific study design.

## Results

### Prescribing durations

Across the five medicines in the basket (i.e. commonly taken once daily for long-term conditions), 160 million prescriptions were issued in England during our 12-month study period (the ten most commonly prescribed quantities shown in Figure S1), of which 133 million were issued for 28, 56 or 84-day durations (Table 2), totalling 5.11 billion tablets/capsules. Of these, 28-day prescriptions accounted for 48.5% (2.5 billion tablets/capsules), 56-day prescriptions accounted for 43.6% (2.2 billion), while 84-day prescriptions accounted for only 8.0% (0.4 billion).

**Table 2.** Total number of items and quantity (tablets/capsules) issued as 28, 56 or 84-day durations across a basket of medicines typically prescribed once daily, in England, Dec 2018-Nov 2019.

| **Quantity per item/ Durations**<br>**(Days)** | **Total Items** | **Total Quantity** | **Proportion of Total Quantity\*** |
| --- | --- | --- | --- |
| **28** | 88,410,515 | 2,475,494,420 | 48.5 |
| **56** | 39,714,191 | 2,223,994,696 | 43.6 |
| **84** | 4,834,340 | 406,084,560 | 8.0 |
| **Total** | 132,959,046 | 5,105,573,676 |  |

*\*Proportions do not sum to 100% due to rounding*

### Geographical variation across England

The 28-day prescribing proportion exhibited wide geographical variation, ranging from 7.2% to 95.0% across England’s 191 CCGs (median 45.6%, interquartile range 28.1% to 65.6%) (Figure 1; Table S1).

**Figure 1.**
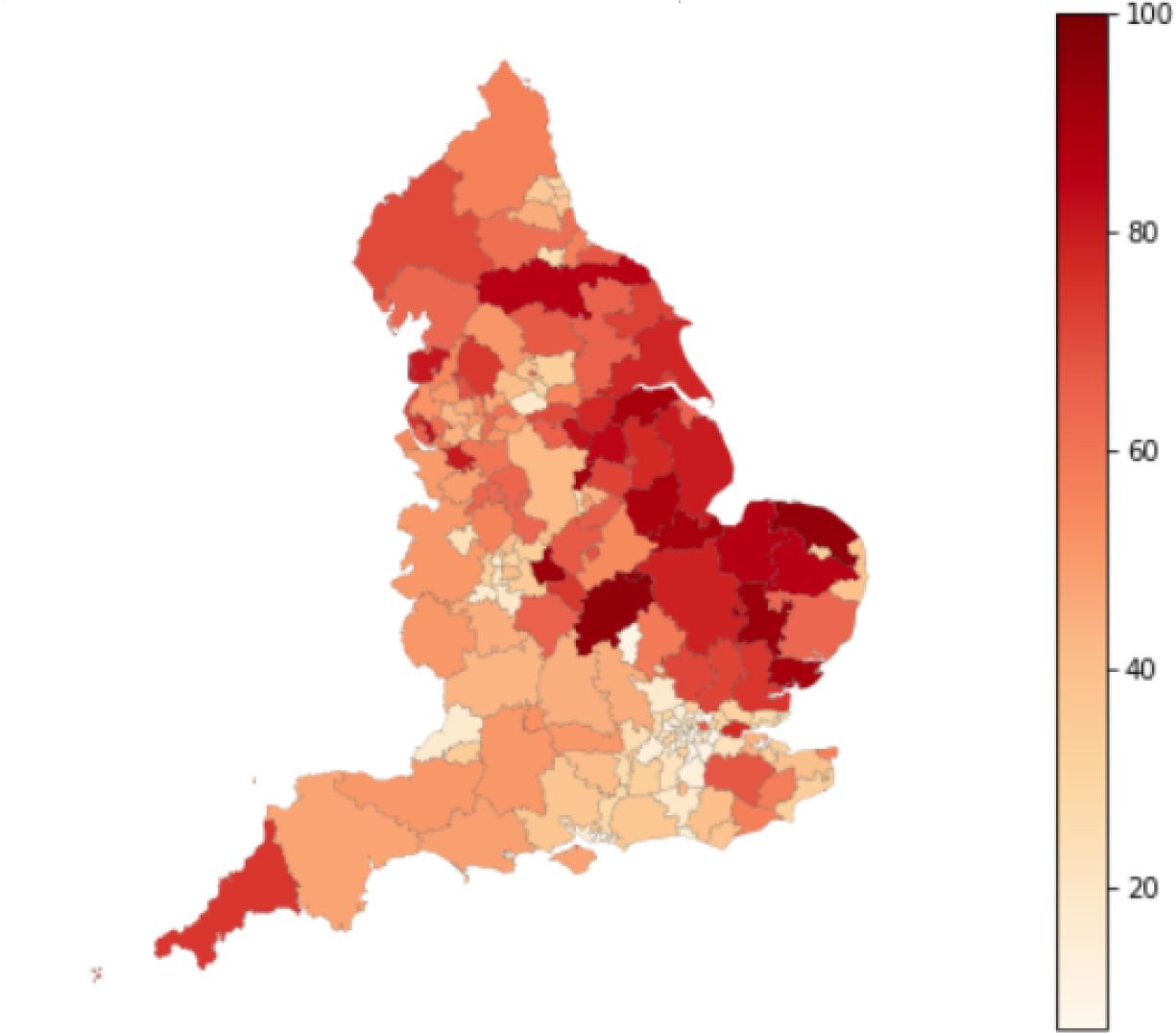
28-day prescribing proportion (%) for the basket of common medicines (Table 1) by each CCG in England. Equivalent maps for 56- and 84-day prescribing proportion (%) are shown in Figure S2.

### Factors associated with prescribing durations

Histograms show 28-day prescribing proportion versus dispensing status (Figure 2), EHR system supplier and LTC quintiles (Figure S3-4).

**Figure 2.**
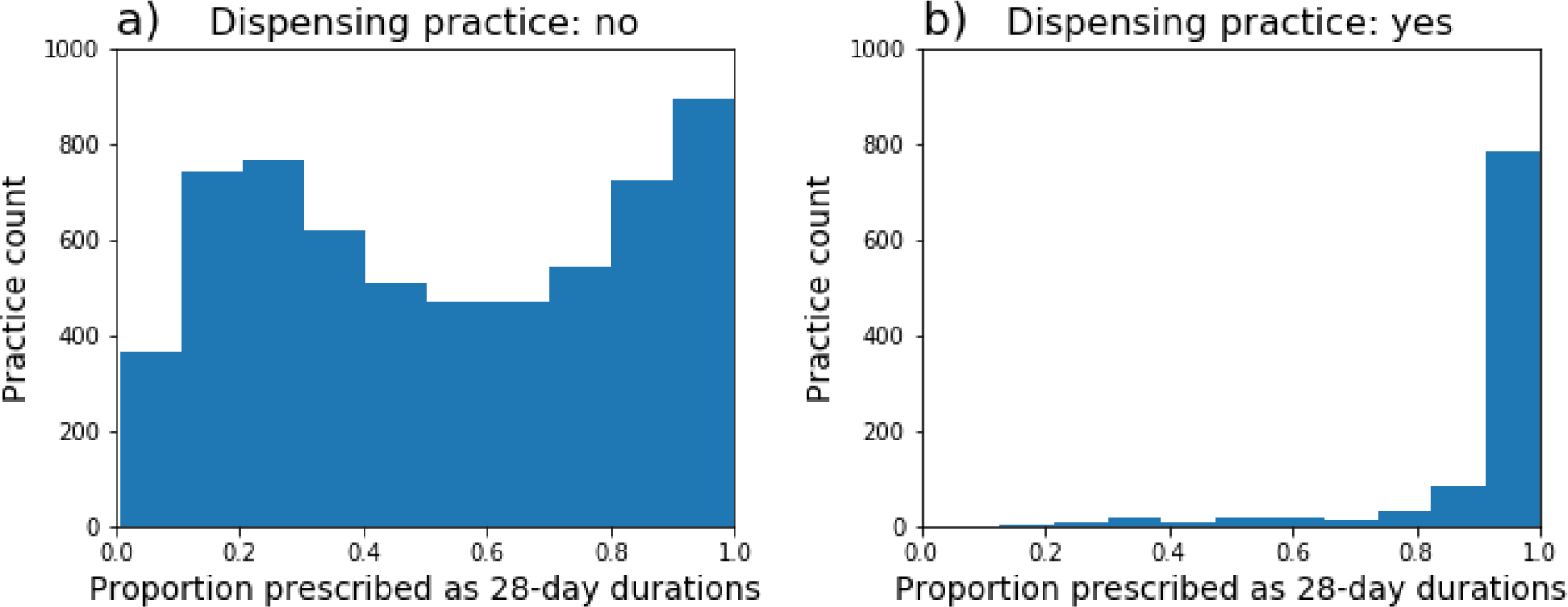
Histograms displaying the distribution of 28-day prescribing proportion for the basket of medicines across all practices in England, split by dispensing status: (a) Non-dispensing, (b) Dispensing, September 2018-August 2019.

The results of the regression analysis showed that each of the investigated factors was significantly associated with the 28-day prescribing proportion, with the exception of the percentage of patients over 65 (Table 3). The strongest association was observed with dispensing status, where dispensing practices had a 64% higher 28-day prescribing proportion than non-dispensing practices (incidence rate ratio (IRR) 1.64, 95% CI 1.49 to 1.80). The percentage of patients with a long-term health condition among the registered population had a dose-response relationship with the 28-day prescribing proportion, with the quintile of practices with the most patients with LTCs prescribing at a 27% higher rate than the lowest quintile (IRR 1.27, 95% CI 1.12 to 1.44). There was also an association with software system, with TPP SystmOne having a higher 28-day prescribing proportion than EMIS (1.11 95% CI 1.02 to 1.21).

**Table 3.** Regression analysis. 28-day prescribing proportion for the basket of medicines (Table 1) vs practice factors.

|  |  | Median proportion 28-day prescribing | Univariable poisson regression |  |  | Multivariable poisson regression |  |  |
| --- | --- | --- | --- | --- | --- | --- | --- | --- |
|  |  |  | Rate ratio | 95% CI |  | Rate ratio | 95% CI |  |
| % of patients over 65 | 0-10.8 | 44.36 | Ref |  |  | Ref |  |  |
|  | 10.8-15.4 | 55.38 | 1.10 | 0.99 | 1.23 | 0.97 | 0.86 | 1.09 |
|  | 15.4-18.9 | 58.21 | 1.18 | 1.06 | 1.30 | 0.92 | 0.81 | 1.05 |
|  | 18.9-22.7 | 62.89 | 1.23 | 1.11 | 1.37 | 0.90 | 0.79 | 1.02 |
|  | 22.7-89.8 | 85.81 | 1.46 | 1.33 | 1.62 | 0.94 | 0.82 | 1.08 |
| % with a long term health condition | 10.0-43.8 | 36.55 | Ref |  |  | Ref |  |  |
|  | 43.8-49.4 | 51.31 | 1.19 | 1.07 | 1.33 | 1.09 | 0.97 | 1.23 |
|  | 49.4-53.6 | 65.07 | 1.34 | 1.21 | 1.49 | 1.17 | 1.03 | 1.32 |
|  | 53.6-58.3 | 70.22 | 1.42 | 1.28 | 1.58 | 1.20 | 1.06 | 1.36 |
|  | 58.3-92.5 | 75.36 | 1.50 | 1.36 | 1.66 | 1.27 | 1.12 | 1.44 |
| EHR system | EMIS | 54.11 | Ref |  |  | Ref |  |  |
|  | Microtest | 93.51 | 1.40 | 0.97 | 2.03 | 1.05 | 0.65 | 1.69 |
|  | TPP SystmOne | 70.27 | 1.15 | 1.08 | 1.23 | 1.11 | 1.02 | 1.21 |
|  | Vision | 55.15 | 0.98 | 0.83 | 1.16 | 1.01 | 0.83 | 1.23 |
| Dispensing status | Not dispensing | 58.91 | Ref |  |  | Ref |  |  |
|  | Dispensing | 96.32 | 1.58 | 1.47 | 1.70 | 1.64 | 1.49 | 1.80 |

*EHR = Electronic Health Record*

### EHR System User-Interface Evaluation

When issuing prescriptions we found that defaults quantities were available for issuing (although can be easily ignored) and that these defaults appeared configurable. Software vendors confirmed that defaults can be configured locally and that when initially implemented they followed the NHS drug tariff which determines community pharmacy reimbursement.

## Discussion

### Summary

Across a small basket of common medications taken once daily, almost half (48.5%) of tablets prescribed over a one-year period in England were in 28-day durations, 43.6% in 56-day (approximately two months) and only 8.0% in 84-day (approximately three months) durations. There was very wide geographic variation (range 7.2% to 95.0% 28-day prescribing proportion across CCGs). Dispensing status was the strongest predictor of short prescription duration. The proportion of patients with LTCs and the EHR software used by a practice were also associated with prescription duration.

### Strengths and weaknesses

Our data includes all prescribing in all typical practices in England, thus minimising the potential for obtaining a biased sample. We used real prescribing data which are sourced from pharmacy claims and therefore did not need to rely on surrogate measures. We excluded a small number of settings such as walk-in centres, which typically do not issue repeat prescriptions for medicines. The data does not currently include dosage instructions on durations, so our findings were limited to medicines typically issued as once-per-day tablets/capsules. Our data only includes data up to 2019. The manuscript development and submission was delayed by the outbreak of the COVID-19 pandemic.

### Findings in Context

To the authors’ knowledge this article represents the first research using large-scale national data to estimate repeat prescription durations. Payne et al studies found that 3-month repeat prescriptions may be more cost effective and suggested that the NHS should encourage extension of repeat prescriptions from one month to three months. Our analysis indicates that less than 50% of prescriptions for commonly prescribed once a day medications are for one-month duration.

A practices dispensing status had the strongest association with prescription duration, with dispensing practices almost exclusively prescribing 28-day courses for this basket of medicines. A potential explanation of this association is that dispensing practices are incentivised to prescribe medication with 28-day duration due to the volume-based dispensing remuneration payment in place. We previously found that dispensing practices were more likely to prescribing higher cost drugs, where they may negotiate lower prices while being reimbursed at a standard rate.[17] Another reason shorter durations may be preferred by dispensing doctors is greater adherence to policies which have endorsed 28-day prescribing in local areas or for stock inventory management e.g. more storage space would be needed to hold stock to supply longer durations.

The percentage of patients with LTCs was also associated with prescription duration. The more patients in a practice have LTCs, the more will have multiple LTCs, likely requiring multiple treatments and more frequent monitoring and/or medication adjustments. Some medications have restrictions on their prescribed duration (e.g. Schedule 2 controlled drugs legally restricted to a 30-day supply) and the prescription duration for all co-prescribed medications is often determined by the one with the shortest acceptable duration (to avoid ordering synchronisation issues) so not all 28-day prescriptions will be suitable to extending to two or three monthly. Additionally for certain clinical conditions, medications [18] and individual patient circumstances, it will be necessary to have some flexibility in policy implementation.

We also found that the EHR used by a practice was associated with prescription duration, most notably TPP had a higher 28-day prescribing proportion than EMIS. We found that both the EMIS and TPP prescribing interfaces used the drug tariff recommended duration as the default (28-days for our basket of medicines), but this could be locally configured if desired by a local primary care organisation in line with their repeat prescription policy. Our previous work has shown that the user interface of each EHR can substantially affect prescribing quality, safety and cost.[19–22] As EHR system of choice is geographically clustered, it is possible that this effect may be driven by adherence by GPs to policies of primary care organisations unrelated to the interface of the EHR.

### Policy implications

A recent report by the Chief Pharmaceutical Officer for the NHS to the government on overprescribing, identified that national organisations should support practices to improve consistency of repeat prescribing. Our analysis clearly indicates inconsistency in repeat prescription duration across England, identifies opportunities for intervention at a policy level as well as providing data to inform the policy debate on current practice [23]

Payne et al identified potential resource savings and our analysis identified that 49% are currently prescribed monthly. Accepting the underlying assumption of colleagues from Bristol, this is likely to save time for clinicians, time for patients’ interactions with the health service in addition to broader benefits like reduced carbon emissions from trips to and from the pharmacy. However we caution that substantial cash releasing savings are unlikely to be reasonable within the current policy context. The same amount of medications need to be purchased by the NHS, although a small amount of savings through reduced wastage is possible. Dispensers (both community pharmacies and dispensing doctors) are paid on a per item basis and if there was a substantial shift to longer prescriptions it would likely lead to reduced income with minimal work load reductions, however our data also shows substantial variation in current patterns suggesting the status quo may not be optimal or indeed fair to dispensers. We show that current reimbursement structures may incentivise shorter prescription durations and that reimbursement varies unevenly across the country with limited regard to patient preference and propose that our data can assist with repeat prescription policy development to provide a more consistent and equitable repeat prescribing experience for patients.

### Future Research

This study sets the foundations for an economic evaluation based on real-world evidence using OpenPrescribing.net and potentially patient level GP data linked to outcome data available via secure platforms like OpenSAFELY, subject to appropriate permissions. An analysis using OpenSAFELY could provide information about health outcomes and NHS resource use associated with duration prescriptions for LTCs in the long term. A targeted emulation trial may be a good alternative to expensive and long Randomised Control Trials to harness the full potentials of using such large datasets to estimate the potential positive (e.g. reduced dispense fees, convenience and saved time of patients and prescribers) and negative (e.g. reduced benefits of medication review/monitoring, medication waste) consequences of extending prescription duration from 28 days to either 56 or 84 days. This is in line with the call of Payne et al for better designed evaluation studies on this topic in the UK that use EHRs to facilitate data collection in near real time which can be analysed by platforms such as OpenSAFELY. Such an emulated targeted trial could facilitate a pragmatic, unobtrusive and low cost evaluation across the whole country. Besides the traditional cost-per-QALY estimates, such an evaluation should provide evidence about potential impact on health inequalities and patient experience as they are both high in the NHS policy mandates. In addition, capacity issues should also be investigated considering the constrained resources in GP practices. The multi-composite evidence of such an evaluation could support the implementation of new repeat prescription policies. We are now seeking collaborators and resources to undertake this analysis.

### Conclusion

One month prescriptions are common for patients taking medicines routinely for long term conditions, particularly in dispensing practices. Electronic health records offer an opportunity to implement and evaluate new policies on repeat prescription duration in England.

## Data Availability

Data management was performed using Python 3 and Google BigQuery, with analysis carried out using Stata 13.2 / Python 3. Data is openly available from the NHS and all code for data management and analysis are archived online.[16]

https://github.com/ebmdatalab/Rx-Quantity-for-Long-Term-Conditions

## Administrative

### Funding

This work was supported by The NIHR Biomedical Research Centre, Oxford, A Health Foundation grant (Award Reference Number 7599); A National Institute for Health Research (NIHR) School of Primary Care Research (SPCR) grant (Award Reference Number 327); the National Institute for Health Research (NIHR) under its Research for Patient Benefit (RfPB) Programme (Grant Reference Number PB-PG-0418-20036) and by the National Institute for Health Research Applied Research Collaboration Oxford and Thames Valley. The views expressed in this publication are those of the author(s) and not necessarily those of the NIHR, NHS England or the Department of Health and Social Care. Funders had no role in the study design, collection, analysis, and interpretation of data; in the writing of the report; and in the decision to submit the article for publication.

### Ethical approval

This study uses exclusively open, publicly available data, therefore no ethical approval was required.

### Guarantor

HC is guarantor.

### Contributorship

BMK, HC and AW conceived the study and designed the methods with input from BG. HC, AW and BMK collected and analysed the data with methodological and interpretation input from AB and BG. HC, AB and BMK drafted the manuscript. All authors contributed to and approved the final manuscript.

