## Supplemental All for "Variation in prescription duration for long term conditions: a cohort study in English NHS primary care using OpenPrescribing" (from another version of this preprint: version 1, 2024-05-29)

### Supplementary Information

***Table S1.*** *Summary statistics describing the 28-, 56- and 84-day prescribing proportion (%) for the basket of common medicines (Table 1) across all Clinical Commissioning Groups (CCGs) in England, December 2018-November 2019.*

|  | **Quantity per item/ Durations (Days)** | | |
| --- | --- | --- | --- |
|  | **28** | **56** | **84** |
| **count** | 191 | 191 | 191 |
| **mean** | 47.2 | 44.1 | 8.7 |
| **std** | 23.4 | 19.4 | 8.2 |
| **min** | 7.2 | 3.8 | 1.0 |
| **25%** | 28.1 | 29.9 | 3.3 |
| **50%** | 45.7 | 45.1 | 5.9 |
| **75%** | 65.6 | 59.4 | 10.9 |
| **max** | 95.0 | 83.2 | 44.2 |

*std = standard deviation*

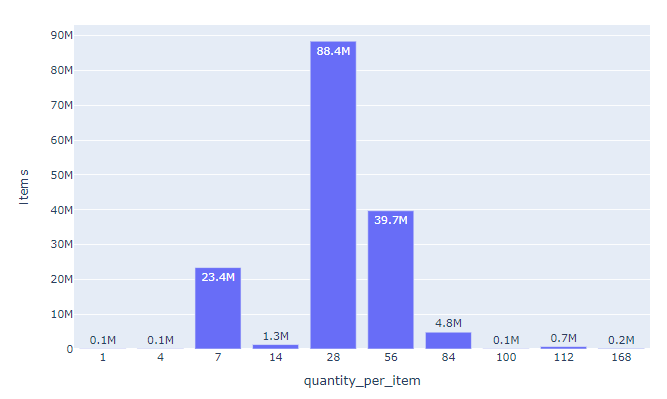

*Figure S1. Histogram displaying the number of items (prescriptions) issued across a basket of medicines typically prescribed once daily, for the ten most commonly prescribed quantity-per-item (equivalent to prescription duration in days), in England Dec 2018-Nov 2019.*

**a)**

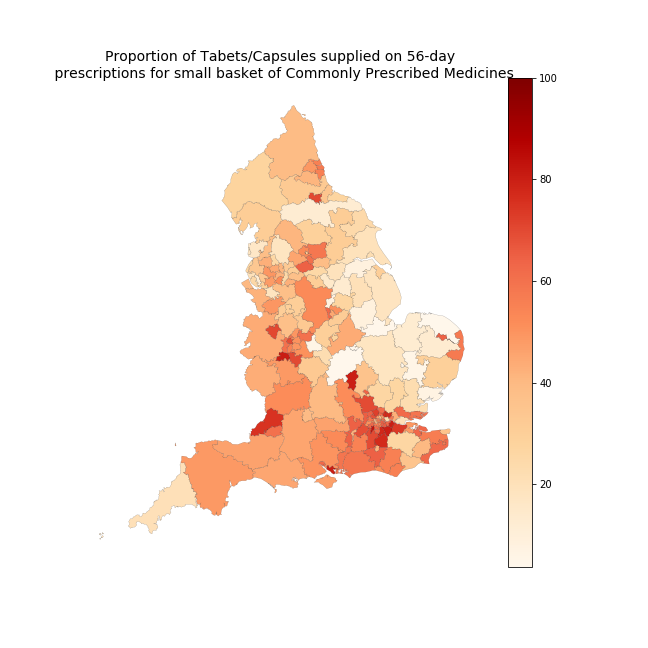

**b)**

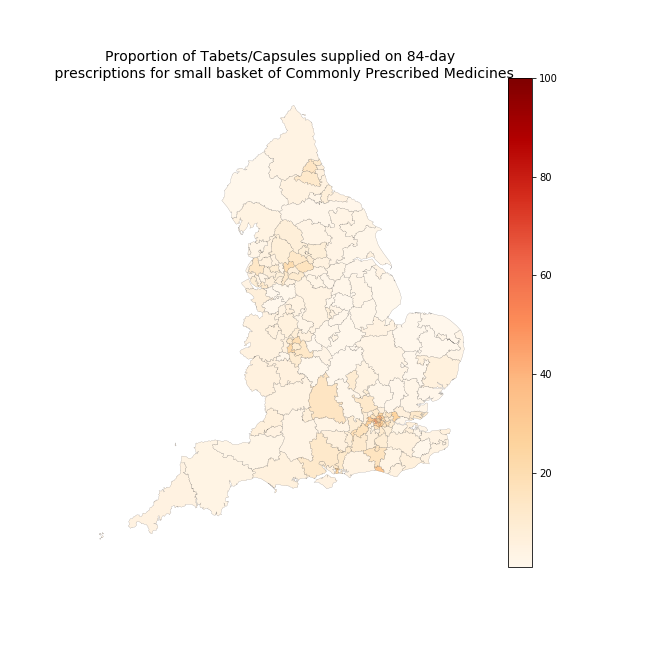

***Figure S2.*** *(a) 56- and (b) 84-day prescribing proportion (%) for the basket of common medicines (Table 1) by each CCG in England.*

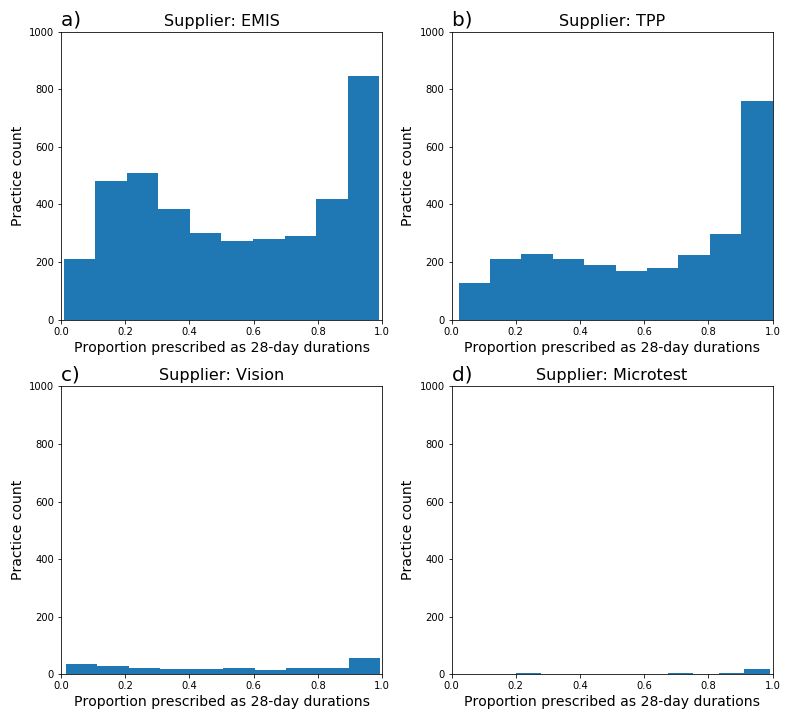

***Figure S3****. Practice counts vs 28-day prescribing proportion for the basket of medicines across all practices in England, split by EHR system supplier: (a) EMIS (b) TPP SystmOne, (c) Vision, (d) Microtest; September 2018-August 2019.*

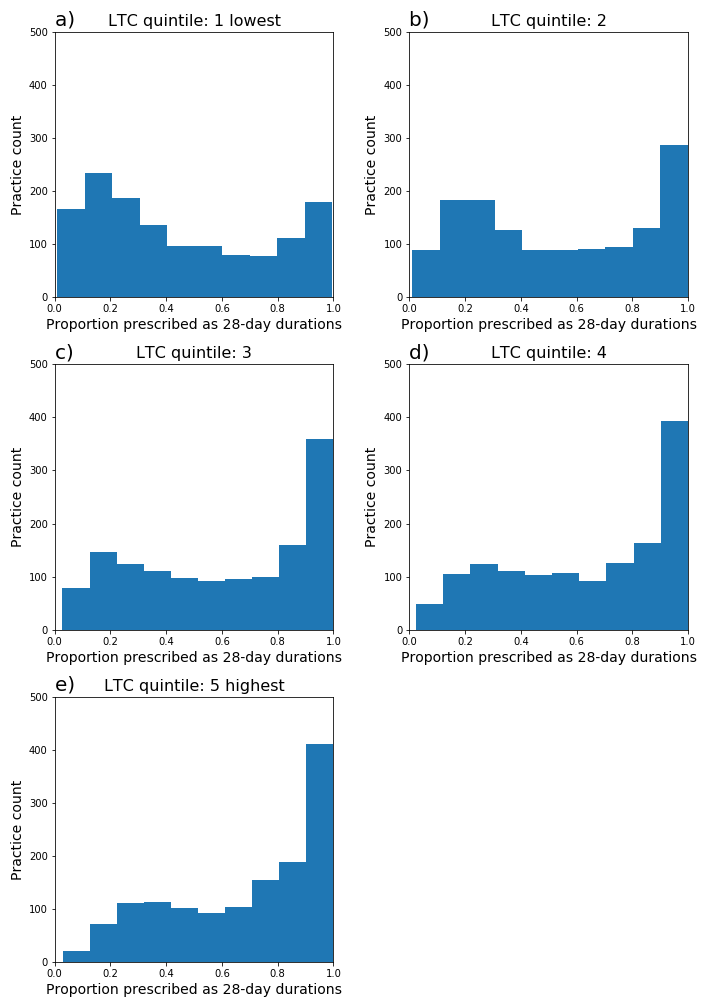

***Figure S4****. Practice counts vs 28-day prescribing proportion for the basket of medicines across all practices in England, split by percentage of patients with an LTC as quintiles, from (a) practices with the lowest percentage of patients with an LTC to (e) those with the most; September 2018-August 2019.*
